# Remote cognitive tests predict neurodegenerative biomarkers in the Insight 46 cohort

**DOI:** 10.1101/2024.09.28.24314472

**Authors:** Martina Del Giovane, Valentina Giunchiglia, Ziyuan Cai, Marguerite Leoni, Rebecca Street, Kirsty Lu, Andrew Wong, Maria Popham, Jennifer M. Nicholas, William Trender, Peter J. Helleyer, Thomas D. Parker, Heidi Murray-Smith, Paresh A. Malhotra, Sebastian J. Crutch, Marcus Richards, Adam Hampshire, Jonathan M. Schott

**Affiliations:** Imperial College London, Department of Brain Sciences. Burlington Danes, the Hammersmith Hospital, 72 Du Cane Road, London, England, W12 0NN, United Kingdom; Imperial College London and the University of Surrey, UK Dementia Research Institute Care Research and Technology Centre. Sir Michael Uren Hub, 9^th^ Floor, 86 Wood Lane, London, England, W12 0BZ, United Kingdom; Department of Neurodegenerative Disease, The Dementia Research Centre, UCL Queen Square Institute of Neurology, London, England, WC1N 3BG, United Kingdom; MRC Unit for Lifelong Health and Ageing at UCL, University College London, 1-19 Torrington Place, London, England, WC1E 7HB, United Kingdom; Department of Medical Statistics, London School of Hygiene and Tropical Medicine, Keppel Street, London, England, WC1E 7HT, United Kingdom; Department of Neuroimaging, Institute of Psychiatry, Psychology and Neuroscience, King’s College London, De Crespigny Park, London, England, SE5 8AF, United Kingdom; Department of Biomedical Informatics, Harvard Medical School, 10 Shattuck St, Boston, MA, 02115, USA

## Abstract

**BACKGROUND:** Alzheimer’s disease-related biomarkers detect pathology years before symptoms emerge, when disease-modifying therapies might be most beneficial. Remote cognitive testing provides a means of assessing early changes. We explored the relationship between neurodegenerative biomarkers and cognition in cognitively normal individuals.

**METHODS:** We remotely deployed 13 computerised Cognitron tasks in 255 Insight 46 participants. We generated whole brain, hippocampal, and white matter hyperintensity volumes at ages 69-71, rates of change over two-years, amyloid load and positivity. We examined the relationship between Cognitron, biomarkers, and standard neuropsychological tests.

**RESULTS:** Slower response time on a delayed recognition task predicted amyloid positivity (OR=1.79,CI:1.15, 2.95). Brain and hippocampal atrophy rates correlated with poorer visuospatial performance (*b*=-0.42, CI:-0.80, -0.05) and accuracy on immediate recognition (*b*=-0.01, CI:-0.01, -0.001), respectively. Standard tests correlated with Cognitron composites (rho=0.43, p<0.001).

**DISCUSSION:** Remote computerised testing correlates with standard supervised assessments and holds potential for studying early cognitive changes associated with neurodegeneration.

## 1 Background

The pathological changes of Alzheimer’s disease (AD) begin years before symptoms emerge [1–3]. Recent trials of anti-amyloid therapies showed beneficial effects in individuals with earlier symptomatic disease [4,5] and these medications are now being trialled in asymptomatic individuals with AD pathology [6]. However, these therapies come with significant risks 4,5], and not all individuals with AD pathology will develop cognitive decline [7]. Therefore, there is a pressing need to identify early cognitive signs associated with AD pathology, which could aid in targeting interventions to those most likely to benefit. Additionally, tools sensitive to subtle changes in memory decline are needed as outcome measures for presymptomatic clinical trials.

Among the available biomarkers included in the National Institute on Aging and Alzheimer’s association criteria [8], cerebral β-amyloid (Aβ) deposition, detectable in vivo through Aβ- PET imaging, is considered core to identify AD at both its asymptomatic and symptomatic phase [9]. Magnetic resonance imaging (MRI) provides additional information about the stage and magnitude of the disease. At the early stages of the disease, where the degree of atrophy measured cross-sectionally may not be sufficient to indicate abnormality, techniques that measure the atrophy rate on serial MRI (e.g., the boundary shift integral) can provide a more sensitive measure of neurodegeneration [10,11].Vascular burden quantified on MRI as white matter hyperintensity volume (WMHV) can be used as a marker of co-pathology. Increased WMHV is predictive of the downstream neurodegeneration in the early symptomatic stage of AD and shorter time to AD development [12,13].

It is challenging to distinguish the earliest cognitive impairments emerging from AD pathology from those associated with normal aging. These impairments may be too subtle, domain-specific, or influenced by demographic factors to be easily detected using standard on-paper assessments [14,15]. Computerised cognitive assessment offers advantages, providing simultaneous measurement of multiple behavioural measures and reaction time, and greater precision. Computerised testing can be scaled for difficulty and complexity to target cognitive impairments in the clinical population of interest. This enhances sensitivity and has the potential to reduce the sample size required for appropriate statistical power in clinical trials involving individuals at the early stage of AD [16,17]. Computerised testing can also be deployed longitudinally with higher stimuli variability and reduced learning effects. To maximize feasibility and sensitivity, tasks should be brief, easy to perform, targeted to cognitive domains thought to be affected in AD, and validated against gold-standard assessments.

We remotely deployed 13 computerised cognitive tasks in the Online 46 study, the remote cognitive sub-study of the MRC National Survey of Health and Development (NSHD) study, a population-based cohort of individuals born in England, Scotland, and Wales in 1946 [18,19]. The aims of this study were to a) identify tasks that were sensitive to markers of AD pathology and neurodegeneration in individuals without dementia; and b) to determine their degree of correlation with standard supervised neuropsychological assessments. The NSHD cohort is particularly suited for the scope of this study. At their current age, the study members are generally at risk of accumulation of AD pathology, prior to the development of dementia, enabling cognitive measures to be benchmarked in relation to these pre-clinical biological changes [20,21].

Our primary hypothesis was that memory performance would demonstrate significant sensitivity to AD pathology as memory impairment is widely recognised as one of the cognitive hallmarks of AD, emerges early in the disease process, and has previously been shown to related to amyloid deposition in this cohort [22–24]. However, memory impairments may occur in non-AD dementias [25], and amyloid deposition has been reported to impact other domains, such as language, attention, visuospatial abilities, and working memory [25,26]. We therefore took an additional exploratory approach, administering a broad set of cognitive tasks to investigate the relationship of AD and vascular pathology with other cognitive sub-domains.

## 2 Methods

### 2.1 Participants and recruitment

The NSHD cohort includes extensive clinical, physical, and cognitive characterisation of an original group of 5362 individuals born in in England, Scotland, and Wales within the same week of March 1946 (https://nshd.mrc.ac.uk/). Online 46 is a remote cognitive sub-study of the NSHD designed to assess the feasibility and utility of remote computerised cognitive testing in this cohort. Online 46 took place between June and September 2023 and included a final sample of 813 participants who completed a battery of 13 cognitive tasks within 4 weeks of invitation. Participants were invited via email and asked to complete the tasks under unsupervised conditions using any electronic device (e.g., tablets, phones, computers) and web browser. Written instructions were provided at the beginning of each task, followed by a set of practice trials to confirm that the participants properly understood how to complete the tasks.

Here we present data from 274 participants who undertook the Online 46 remote cognitive battery and were also part of the Insight 46 neuroimaging sub-study. Insight 46 is a prospective longitudinal observational neuroimaging sub-study of the NSHD. It involves detailed clinical, neuropsychological testing and neuroimaging data, which have been used for the present study. The recruitment procedure is summarised in Figure 1 and the data collection methods for Insight 46 have been described previously described [18,19,27].

**Figure 1.**
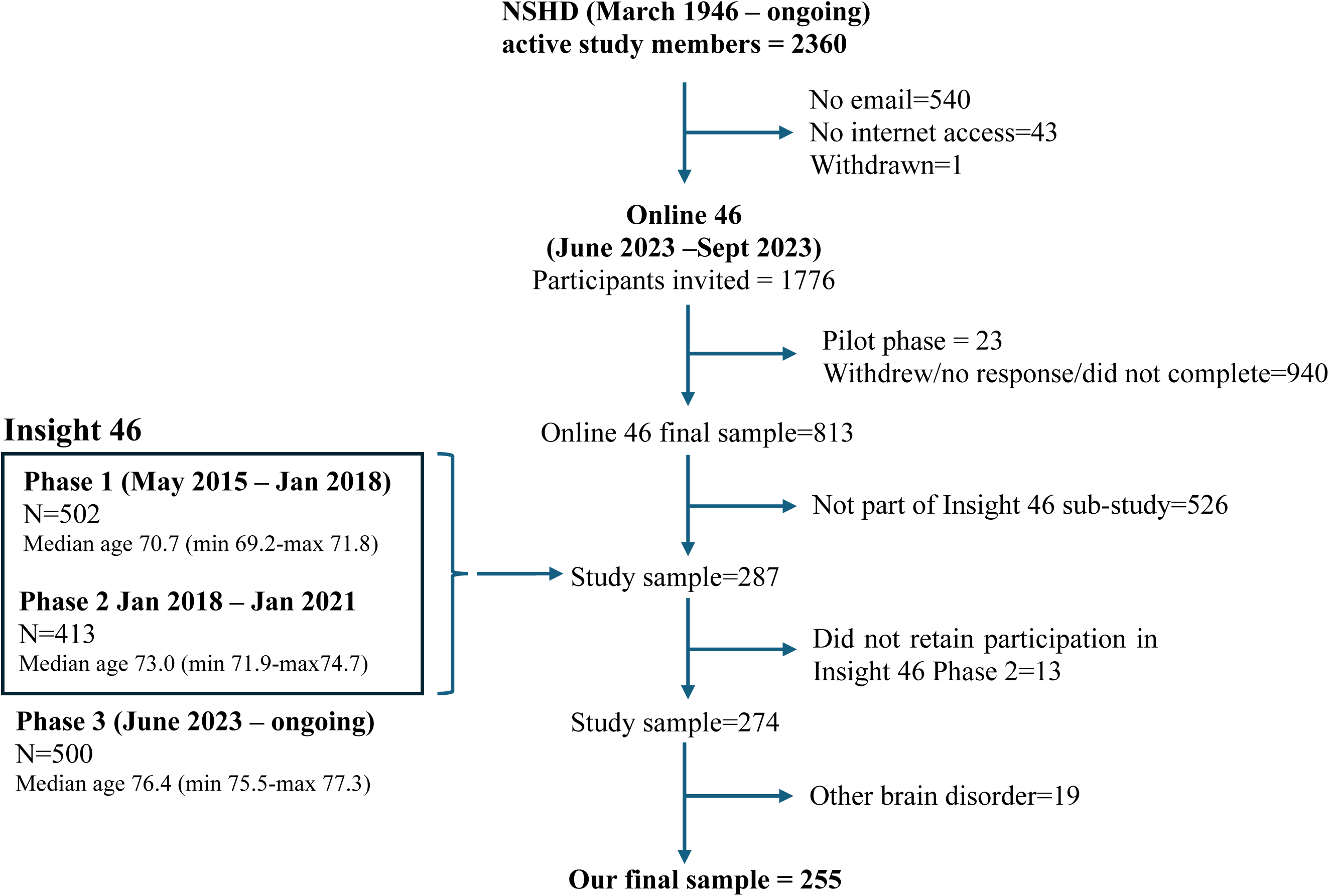
Recruitment flowchart

As our focus was on presymptomatic cognitively normal individuals, participants were excluded if they had evidence of major brain disorders including any clinically diagnosed neurodegenerative disorder, any psychiatric disorder requiring an antipsychotic treatment, depression necessitating electroconvulsive shock therapy, evidence of traumatic brain injury or significant neurosurgery, multiple sclerosis, evidence of an ischemic or haemorrhagic cortical stroke, radiological evidence of a brain malignancy, and mild cognitive impairment (MCI).

### 2.2 Cognitive testing

A library of computerised tasks developed in HTML5 with JavaScript is available on the Cognitron platform (https://www.cognitron.co.uk) [28–33]. A battery of 13 of these tasks was deployed remotely to 813 participants of the Online 46 study group at age 77. Compliance, adherence and usability of the tasks have been studied and reported elsewhere [34]. The tasks were selected to cover multiple cognitive domains and target cognitive abilities thought to be impaired early in AD, while being brief and understandable by the study participants. Task descriptions are in Figure 2 and Table 1. Participants received written instructions and completed a brief sequence of practice trials before starting each task to ensure they understood how to complete them. Average completion time for the whole battery was 39 minutes (mean time per task=4.55 minutes, min=1.73, max=5.01). All tasks yielded a summary score that was accuracy-based (total correct answers) and a secondary score that was the median RT, except for motor control which was measured primarily with an RT score (Table 1). As the testing sessions were conducted under unsupervised conditions, we defined signs of non-compliance for each Cognitron task to exclude participants who did not properly engage. Specifically, one participant was excluded from 2D Manipulations for repetitively clicking in the same location of the screen. Two participants were excluded from Digit Span - one for clicking on another browser page while performing the task and the other for achieving an accuracy score below the established threshold for engagement (located on the left tail of the distribution of accuracy scores across the entire cohort). Three participants were also excluded from Spotter as they showed an accuracy score below the expected threshold for engagement. In a separate analysis examining data from the whole Online 46 group (N=813) and reported elsewhere [34], 9% of individuals showed indicators of lack of compliance with the Choice reaction time (CRT) task. The task was therefore considered unreliable and excluded from further analysis.

**Figure 2.**
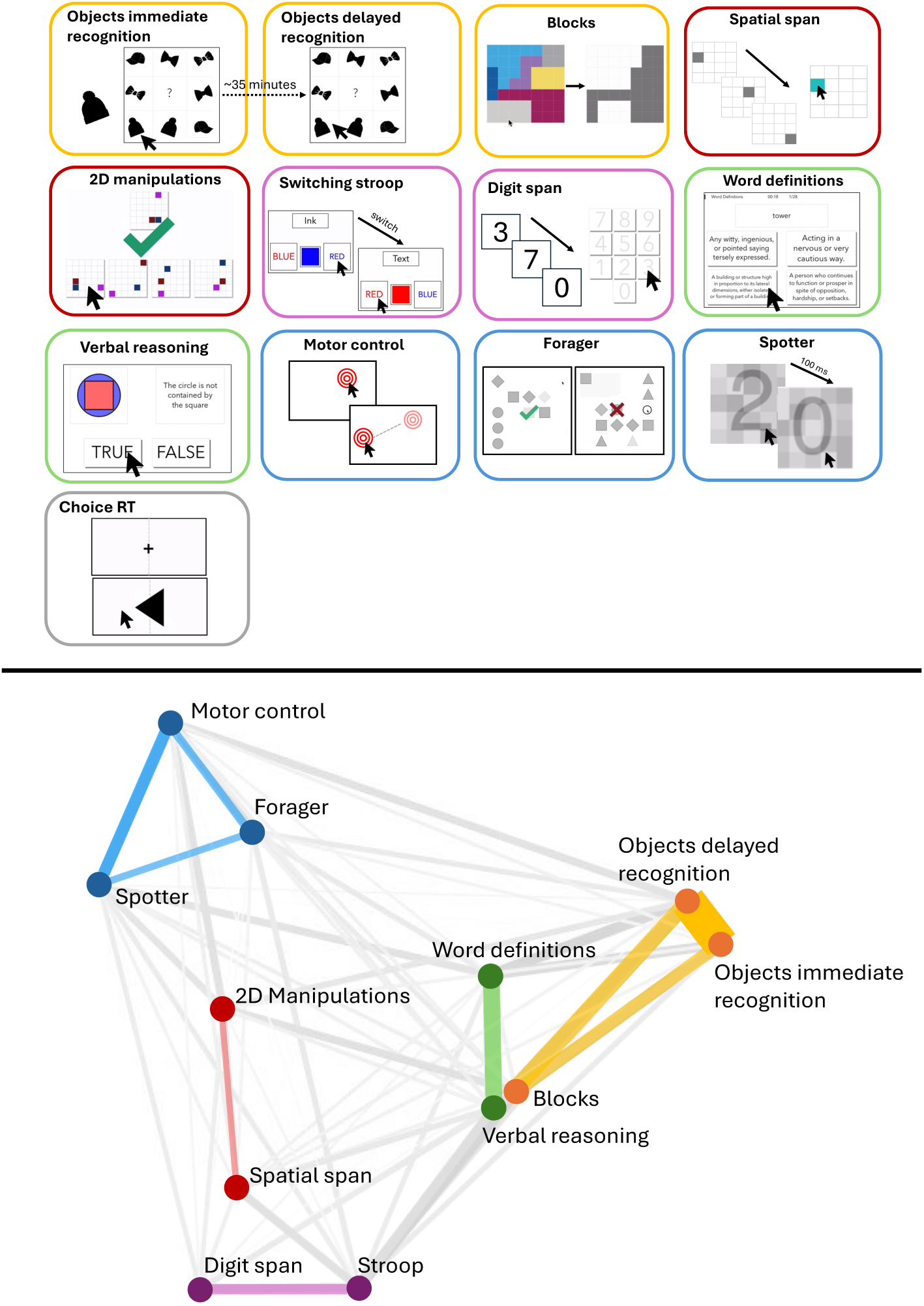
Top: Illustrations of the Cognitron tasks. Bottom: Network plot showing clustering of the primary task scores, and the cognitive domains measured. The colours of the connecting lines indicate the 5 components derived from the PCA. A smaller distance and lower opacity of the connecting line indicate a stronger correlation.

**Table 1.**
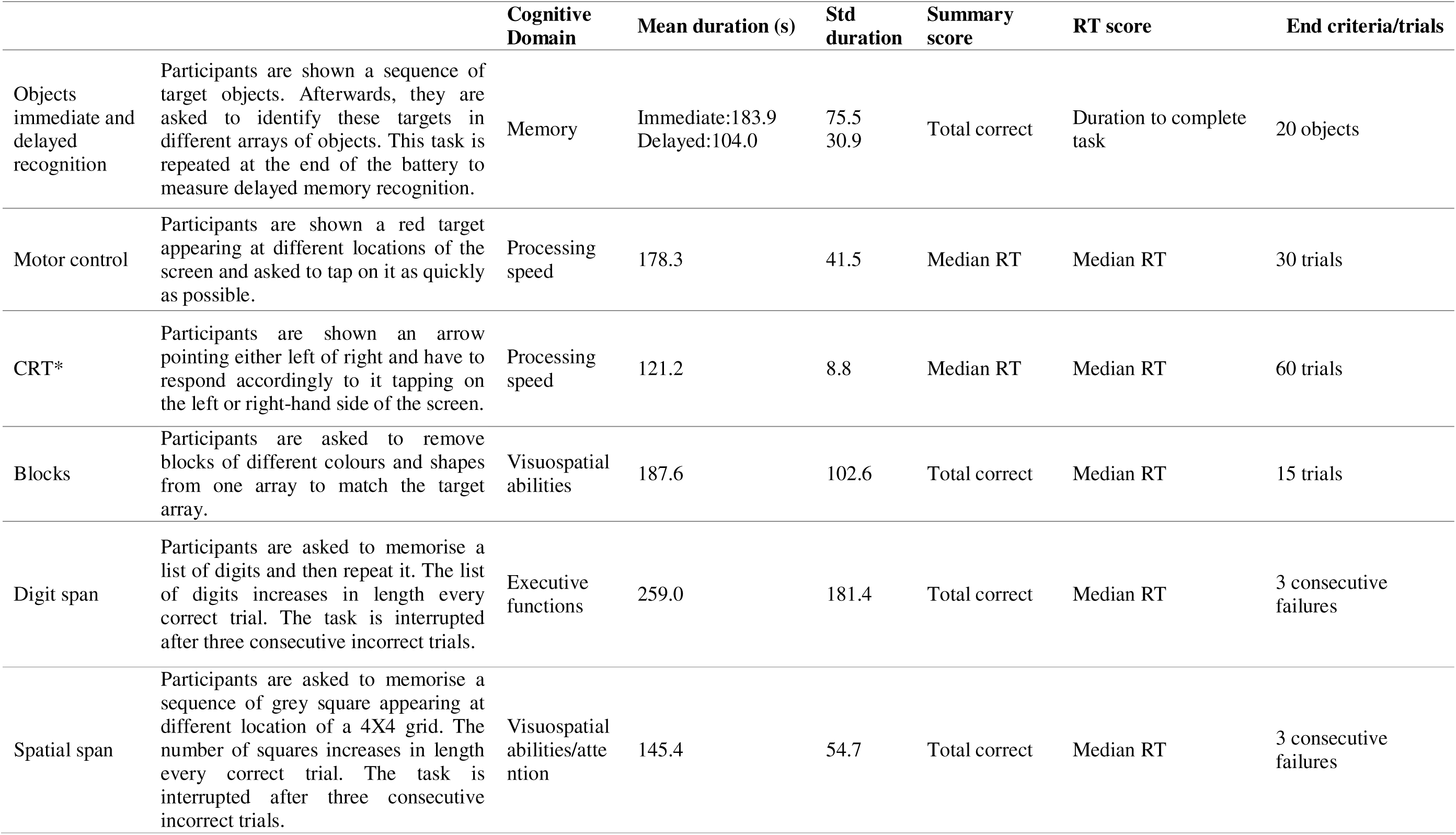
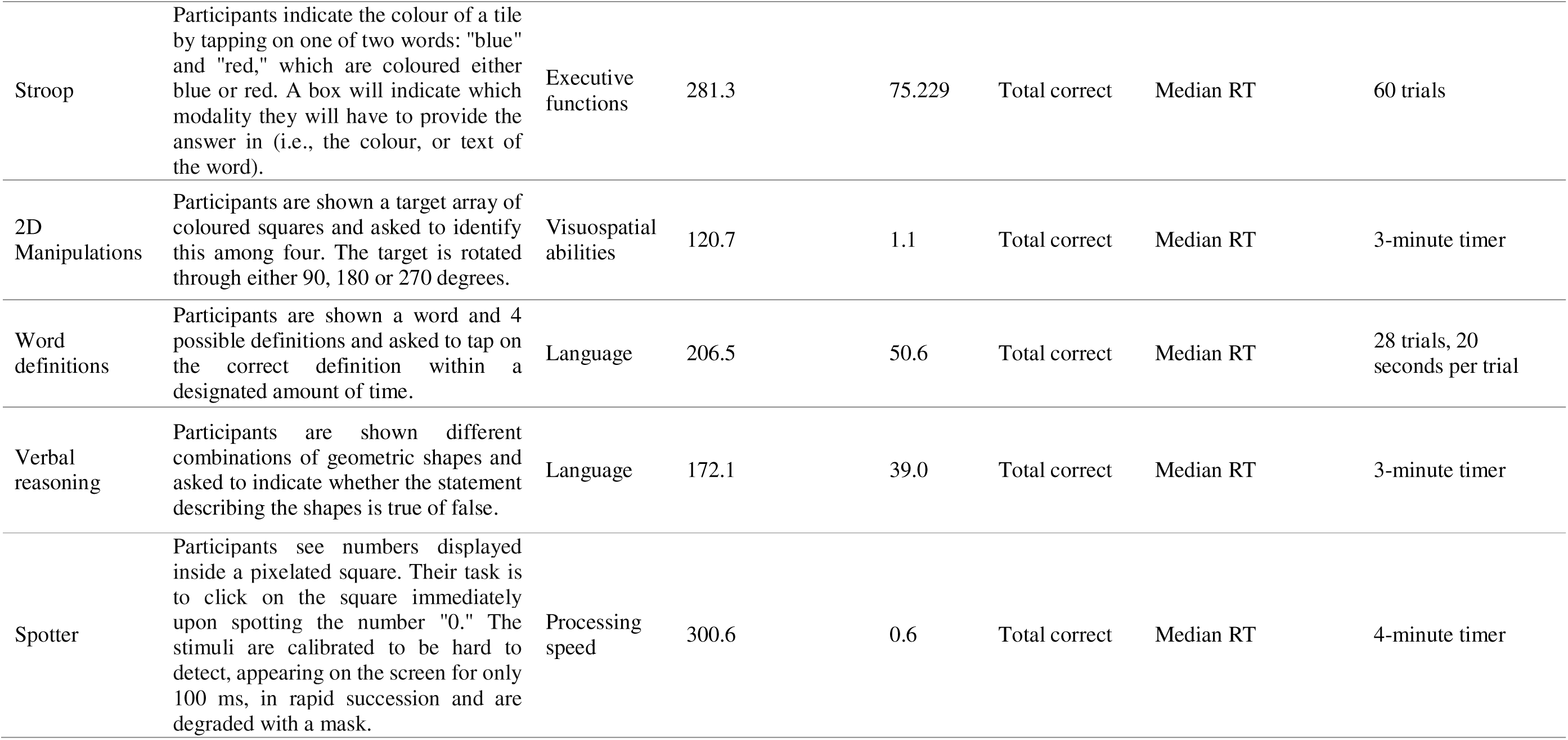
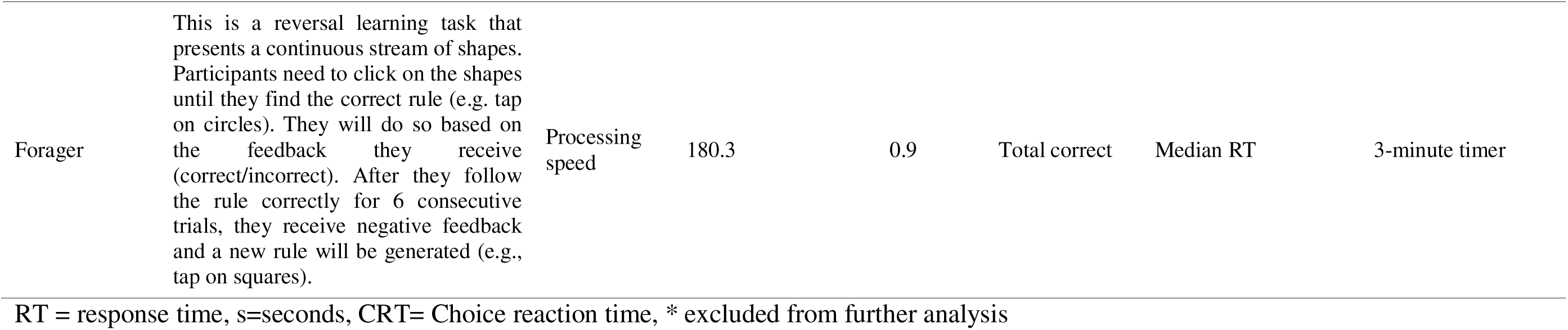
Details of the Cognitron tasks.

|  |  | **Cognitive Domain** | **Mean duration (s)** | **Std duration** | **Summary score** | **RT score** | **End criteria/trials** |
| --- | --- | --- | --- | --- | --- | --- | --- |
| Objects immediate and delayed recognition | Participants are shown a sequence of target objects. Afterwards, they are asked to identify these targets in different arrays of objects. This task is repeated at the end of the battery to measure delayed memory recognition. | Memory | Immediate:183.9<br>Delayed:104.0 | 75.5<br>30.9 | Total correct | Duration to complete task | 20 objects |
| Motor control | Participants are shown a red target appearing at different locations of the screen and asked to tap on it as quickly as possible. | Processing speed | 178.3 | 41.5 | Median RT | Median RT | 30 trials |
| CRT* | Participants are shown an arrow pointing either left or right and have to respond accordingly to it tapping on the left or right-hand side of the screen. | Processing speed | 121.2 | 8.8 | Median RT | Median RT | 60 trials |
| Blocks | Participants are asked to remove blocks of different colours and shapes from one array to match the target array. | Visuospatial abilities | 187.6 | 102.6 | Total correct | Median RT | 15 trials |
| Digit span | Participants are asked to memorise a list of digits and then repeat it. The list of digits increases in length every correct trial. The task is interrupted after three consecutive incorrect trials. | Executive functions | 259.0 | 181.4 | Total correct | Median RT | 3 consecutive failures |
| Spatial span | Participants are asked to memorise a sequence of grey square appearing at different location of a 4X4 grid. The number of squares increases in length every correct trial. The task is interrupted after three consecutive incorrect trials. | Visuospatial abilities/attention | 145.4 | 54.7 | Total correct | Median RT | 3 consecutive failures |
| Stroop | Participants indicate the colour of a tile by tapping on one of two words: "blue" and "red," which are coloured either blue or red. A box will indicate which modality they will have to provide the answer in (i.e., the colour, or text of the word). | Executive functions | 281.3 | 75.229 | Total correct | Median RT | 60 trials |
| 2D Manipulations | Participants are shown a target array of coloured squares and asked to identify this among four. The target is rotated through either 90, 180 or 270 degrees. | Visuospatial abilities | 120.7 | 1.1 | Total correct | Median RT | 3-minute timer |
| Word definitions | Participants are shown a word and 4 possible definitions and asked to tap on the correct definition within a designated amount of time. | Language | 206.5 | 50.6 | Total correct | Median RT | 28 trials, 20 seconds per trial |
| Verbal reasoning | Participants are shown different combinations of geometric shapes and asked to indicate whether the statement describing the shapes is true or false. | Language | 172.1 | 39.0 | Total correct | Median RT | 3-minute timer |
| Spotter | Participants see numbers displayed inside a pixelated square. Their task is to click on the square immediately upon spotting the number "0." The stimuli are calibrated to be hard to detect, appearing on the screen for only 100 ms, in rapid succession and are degraded with a mask. | Processing speed | 300.6 | 0.6 | Total correct | Median RT | 4-minute timer |
| Forager | This is a reversal learning task that presents a continuous stream of shapes. Participants need to click on the shapes until they find the correct rule (e.g. tap on circles). They will do so based on the feedback they receive (correct/incorrect). After they follow the rule correctly for 6 consecutive trials, they receive negative feedback and a new rule will be generated (e.g., tap on squares). | Processing speed | 180.3 | 0.9 | Total correct | Median RT | 3-minute timer |

RT = response time, s=seconds, CRT= Choice reaction time, \* excluded from further analysis

Participants had already completed a comprehensive Insight 46 battery of standard supervised neuropsychological assessments at age 73 [18]. A subset of these standard measures mapping the cognitive domains measured by the Cognitron tasks was selected to study their association with the tasks (Supplementary table 1). These were the Mini-Mental State Examination (MMSE) [35], the Digit Symbol Substitution test (DSST) of the Wechsler Adult Intelligence Scale-Revised (WAIS-R) [36], the Logical Memory test of the Wechsler Memory Scale-Revised (WMS-R) [37], the 12-item Face-Name Associative Memory Exam (FNAME-12) [38], the Preclinical Alzheimer cognitive composite (PACC) generated from the measures listed above [39], and a Trail Making B test [38], the Matrix Reasoning task of the Wechsler Abbreviated Scale of Intelligence (WASI) [40], the Adult Memory and Information Processing Battery’s Complex Figure Drawing task [41], a 15-item word learning task [42], the Graded Naming task [43], and a choice reaction time task [44].

### 2.3 Neuroimaging biomarkers

Participants underwent a 60-minute scan at age ∼69-71 and ∼71-73 on a single Biograph mMR 3T PET/MRI scanner (Siemens Healthcare, Erlangen), with intravenous injection of 370 MBq of 18F-Florbetapir (Amyvid). A/3 burden was assessed during a 10-minute period ∼50 minutes after injection. PET data were processed using an automated pipeline including pseudo-CT (computed tomography) attenuation correction [45]. A global standard uptake value ratio (SUVR) was calculated from a cortical gray matter composite with an eroded subcortical white matter reference region. Positive or negative A/3 status was determined by applying a Gaussian mixture model applied to SUVR values, with the 99th percentile of the A/3-negative Gaussian as the cut-point (0.6104) [23]. Whole brain, hippocampal, and ventricular volume were calculated as previously reported [18]. In cases where data were not available at follow-up (n=28), amyloid status and the extent of amyloid deposition were assessed based on the scans obtained at age 71. The rates of change in whole brain, hippocampal and ventricular volumes between age 71 and 73 were calculated using the boundary shift integral (BSI) [10]. Hippocampal volumes and BSI were derived as the sum of left and right structures. Additionally, white matter hyperintensity volumes were obtained from the MRI scans performed at age 71 using an unsupervised automated algorithm, Bayesian Model Selection to T1 and FLAIR images, generating a global WMHV, which included subcortical grey matter but not infratentorial regions.

### 2.4 Statistical analysis

Demographic differences between amyloid positive and amyloid negative individuals were assessed using Chi-squared tests for gender, handedness and education level, and two sample t-test for childhood cognitive abilities. We then analysed the demographic differences between participants who did and did not attempt the battery of Cognitron tasks. Chi-squared tests were used to examine differences in gender, education, handedness, adult socioeconomic status, amyloid levels, and ApoE status. Two sample t-tests were conducted to assess differences in childhood cognitive abilities and PACC scores. To evaluate differences in MMSE scores, we used the Wilcoxon rank-sum test, as the data was not normally distributed.

We used linear regression to adjust the standard supervised assessments and the Cognitron accuracy and RT scores for demographic characteristics which may influence cognitive performance. Specifically, we included sex, handedness, education level, and the device used to complete the tasks as predictors, and the cognitive scores as dependent variables. Residuals were calculated as the difference between the observed score and the predicted score. These would represent the portion of cognitive performance not explained by the demographic factors. This effectively removes the influence of these variables. The residuals were then scaled by subtracting the mean and dividing by the standard deviation.

We examined the inter-correlations between the Cognitron tasks to confirm that they encompassed distinct cognitive domains. A Principal Component Analysis (PCA) with orthogonal varimax rotation was conducted using the Kaiser convention (eigenvalue > 1). Components were interpreted based on the tasks that had the highest and discrete loadings on them. We then investigated the relationship between the scaled Cognitron scores and biomarkers of neurodegeneration. Multivariable logistic regression was utilised with accuracy and RT measures of Cognitron as predictors and amyloid positivity as dependent variable. A secondary linear regression analysis tested the relationship between the Cognitron task that significantly predicted amyloid status and the extent of amyloid deposition (SUVR). We also ran a linear regression analysis to explore whether the relationship between RT and accuracy on this task differed by amyloid status. We included RT as the independent variable and accuracy as the dependent variable, with amyloid status as an interaction term. We then ran separate linear regression models for each amyloid status group, using RT as the independent variable and accuracy as the dependent variable in each model.

To examine the relationships between the online cognitive scores and MRI variables, linear regression models were conducted inputting the Cognitron accuracy and RT scores as predictors and the volume and BSI for the whole brain, hippocampi, and ventricles, as well as the WMHV as dependent variables. To handle the skewed distribution of WMH volume, generalised linear models with a gamma log link function were employed. All models were conducted both with and without adjustment for childhood cognitive abilities measured at age 8, 11, and 15 using 4 tests of verbal and nonverbal ability devised by the National Foundation for Education Research [46]. We utilised the scores obtained at age 8. In cases where data were not available at this age, we utilised the scores derived at ages 11 or 15. The models including SUVR, whole brain, hippocampal, ventricular and WMH volume as dependent variables were corrected for total intracranial volume (TIV).

Additionally, we looked at the correlation between the Cognitron battery and the battery of standard supervised cognitive assessments. We extracted a composite score from the Cognitron tasks which had shown significant associations with neurodegeneration biomarkers, and a total composite score from the Insight 46 standard supervised assessments. This was done using PCA, where the first principal component was taken as a global composite score. Spearman correlation was used to compare the derived Cognitron and standard assessments total composite scores. The Cognitron composite score was also correlated with the PACC to assess its utility as measure of early AD-related cognitive changes. Finally, we examined the correlation between a memory composite score derived from the Cognitron memory tasks (i.e., Objects Memory Immediate and Delayed Recognition accuracy and RT) and a composite score generated from a subset of Insight 46 standard assessments targeting memory abilities (i.e., Logical Memory, FNAME-12, and AMIPB - Complex Figure Drawing). These composite scores were calculated using PCA, with the first unrotated component serving as the composite memory score. The aim of these analyses was to confirm the diagnostic value of this subset of tasks, comparing their effectiveness to gold-standard assessments, and to inform the development of a more concise Cognitron battery for future studies.

## 3 Results

274 participants undertook the Cognitron battery. Following exclusion of 19 participants with evidence of a brain disorder, 255 individuals were included in the final analysis (51.76% males). Supplementary table 2 illustrates the number of participants included in the analysis for each task. Amyloid status was missing for 14 individuals at an SUVR cut-off of 0.61 [23], and 66 (27.39%) were Aβ-positive. Full details of the demographics characteristics of the sample are shown in Table 2. No significant demographic differences were observed between Aβ-positive and Aβ-negative participants. Of the 122 Online 46 participants who did not attempt the Cognitron battery, 36 (30.5%) were Aβ-positive. Demographic differences between individuals who did and did not attempt the Cognitron battery are reported in Supplementary table 3. In brief, Individuals who did not attempt the battery showed significantly lower childhood cognitive abilities (t_(665.44)_=-6.69, CI:-0.38 to -0.21), and lower performance on the MMSE (W=13229, p<0.001) and PACC (t_(205.76)_=-3.85, CI :-0.44 to - 0.14). The factor analysis indicated that the Cognitron tasks clustered in five interpretable cognitive domains, explaining 61.34% of variance (Figure 2 and 3).

**Figure 3.**
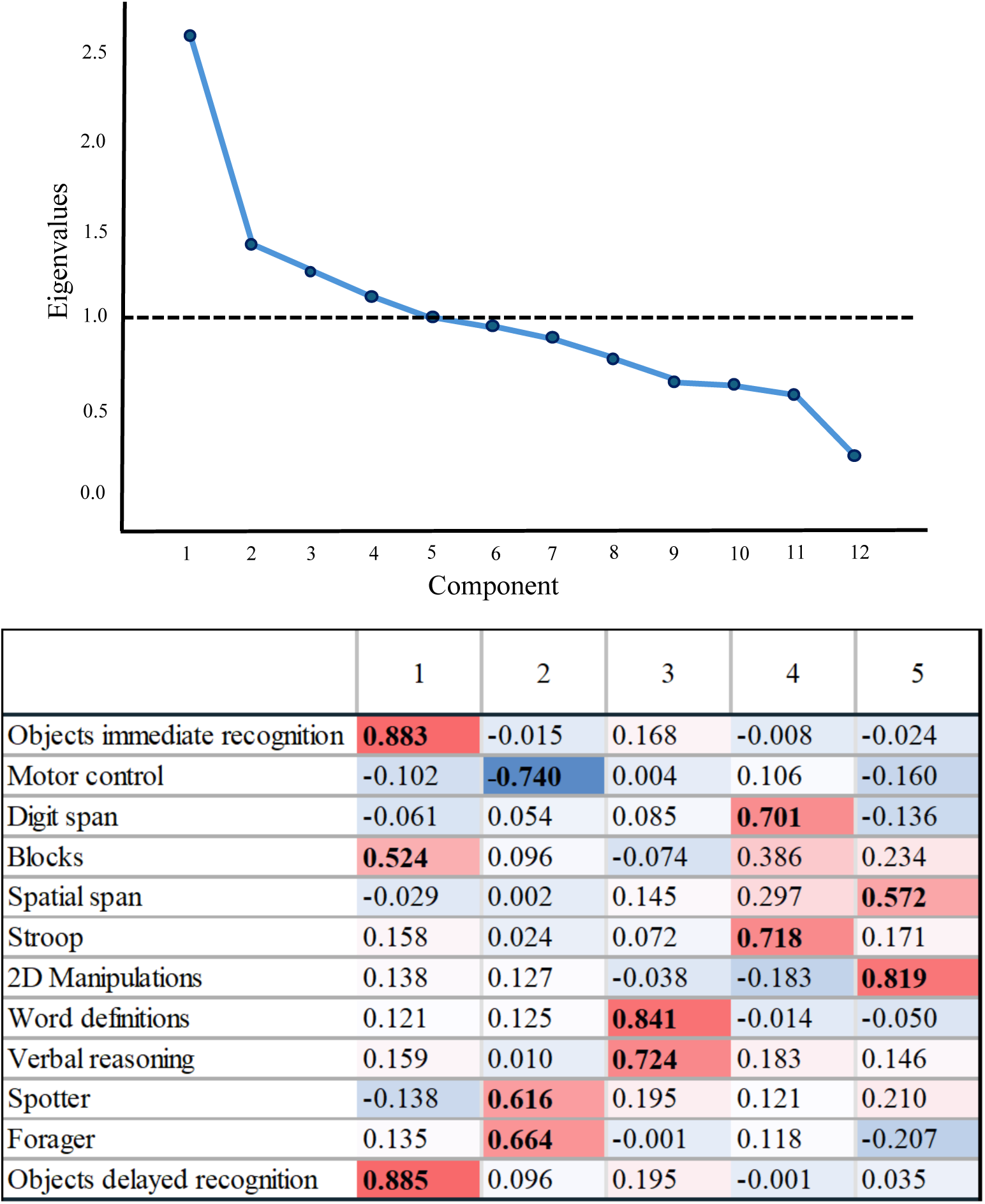
PCA applied to the Cognitron summary scores. Top: Scree plot showing eigenvalues (components extracted based on eigenvalues > 1). Bottom: Loadings of the Cognitron summary scores onto the derived cognitive components

**Table 2.**
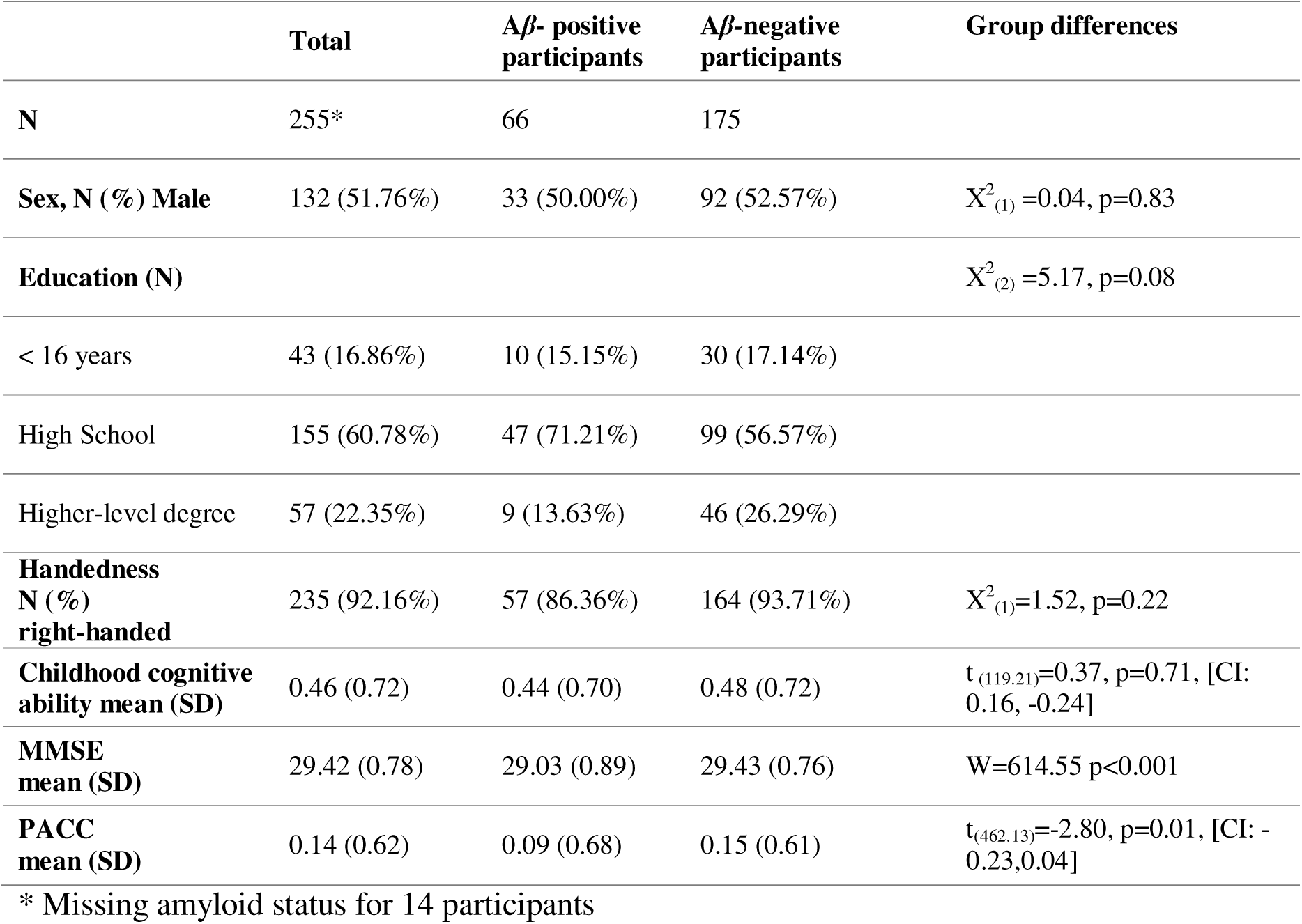
Demographic characteristics of the study groups.

|  | **Total** | **Aβ- positive participants** | **Aβ-negative participants** | **Group differences** |
| --- | --- | --- | --- | --- |
| **N** | 255* | 66 | 175 |  |
| **Sex, N (%) Male** | 132 (51.76%) | 33 (50.00%) | 92 (52.57%) | $X^2_{(1)}=0.04, p=0.83$ |
| **Education (N)** | | | | $X^2_{(2)}=5.17, p=0.08$ |
| < 16 years | 43 (16.86%) | 10 (15.15%) | 30 (17.14%) |  |
| High School | 155 (60.78%) | 47 (71.21%) | 99 (56.57%) |  |
| Higher-level degree | 57 (22.35%) | 9 (13.63%) | 46 (26.29%) |  |
| **Handedness N (%) right-handed** | 235 (92.16%) | 57 (86.36%) | 164 (93.71%) | $X^2_{(1)}=1.52, p=0.22$ |
| **Childhood cognitive ability mean (SD)** | 0.46 (0.72) | 0.44 (0.70) | 0.48 (0.72) | $t_{(119.21)}=0.37, p=0.71, [CI: 0.16, -0.24]$ |
| **MMSE mean (SD)** | 29.42 (0.78) | 29.03 (0.89) | 29.43 (0.76) | $W=614.55 p<0.001$ |
| **PACC mean (SD)** | 0.14 (0.62) | 0.09 (0.68) | 0.15 (0.61) | $t_{(462.13)}=-2.80, p=0.01, [CI: -0.23, 0.04]$ |

\* Missing amyloid status for 14 participants

### 3.1 Associations with neurodegenerative biomarkers

We found that a one-unit increase in RT on the Objects delayed recognition task corresponded to a 1.79-fold increase in the odds of being Aβ-positive (CI: 1.15, 2.95). Conversely, a one-unit increase in accuracy on the same task was associated with a 0.60-fold reduction in the odds of having positive amyloid status (CI: 0.36, 0.99). When examining these associations using amyloid as a continuous measure, we confirmed that higher SUVR was associated with slower RT (0.05, CI: 0.01, 0.10) and reduced accuracy (-0.04, CI:-0.07, - 0.003) on Objects delayed recognition (Figure 4). Using linear regression, we also investigated whether the effect of RT on the accuracy of the Objects delayed recognition task differed in the Aβ-positive and negative groups. We found a significant interaction between RT and amyloid status affecting task accuracy (-0.32, CI: -0.63, -0.01). Further analysis revealed that slower performance was significantly associated with lower accuracy in the Aβ- positive (-0.49, CI:-0.73, -0.25) but not negative (-0.17, CI:-0.38, 0.05) group.

**Figure 4.**
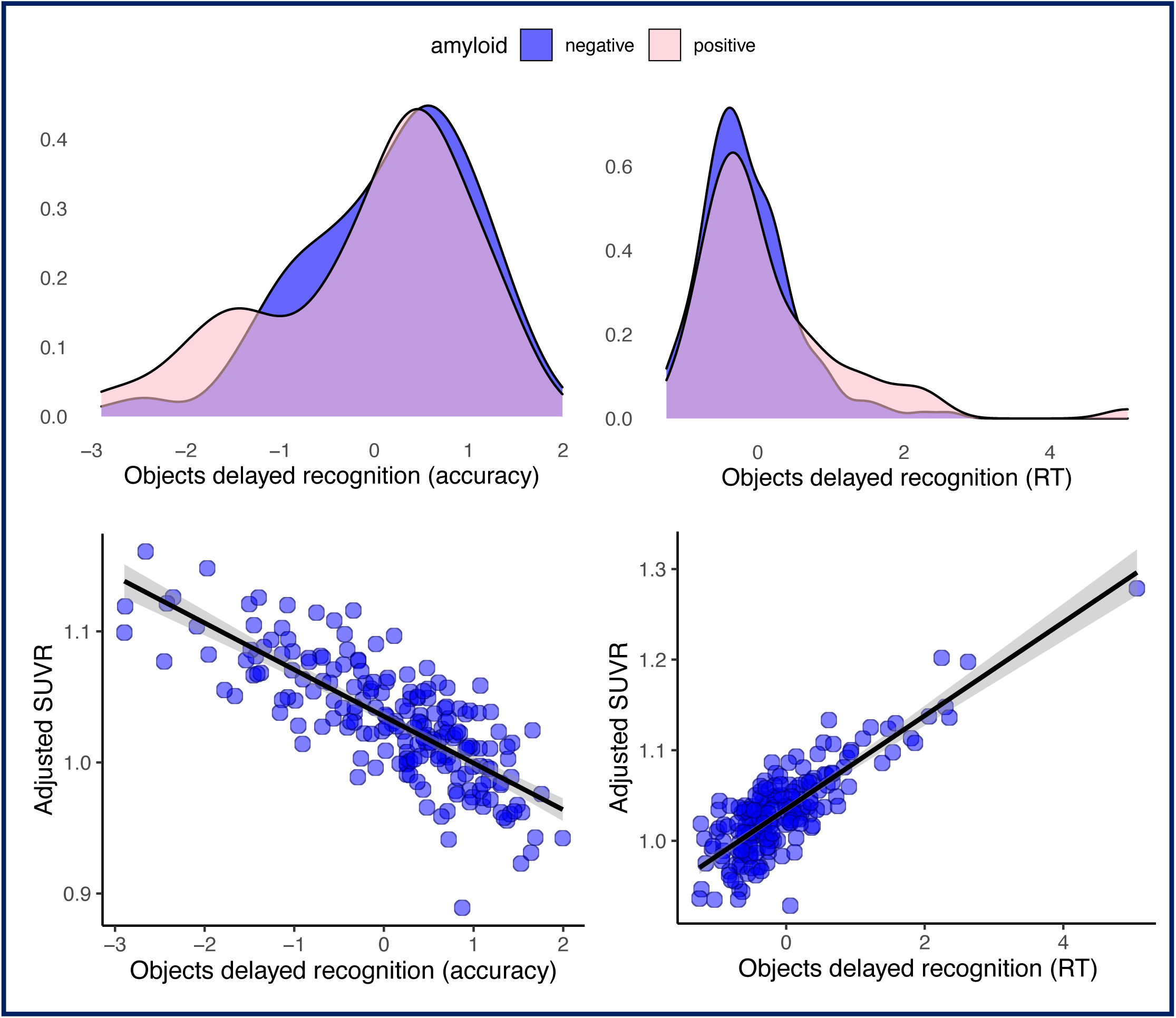
Top: Density plots showing performance differences on the Objects delayed recognition task (accuracy and RT scores) between the amyloid positive and negative groups. Bottom: Association between SUVR and the Objects delayed recognition task (accuracy and RT scores)

The results of the regression models examining the association between the Cognitron tasks and MRI-based biomarkers are presented in Figure 5. A one-unit decrease in accuracy on the Objects immediate recognition task was associated with a 0.01 mL/year faster rate of hippocampal atrophy rate (CI: 0.001, 0.012). Additionally, for each unit increase in RT on the Objects delayed recognition task, the WMHV increased by 1.19 mL (CI: 1.04, 1.57). We also found that a one-unit decrease in accuracy on the Spatial span task corresponded to a 0.42 mL/year increase in whole brain atrophy rate (CI: 0.05, 0.80) and a 0.83 mL increase in cross-sectional WMHV (CI: 0.71, 0.96). Furthermore, each unit increase in RT on the Spotter task corresponded to a 1.19 mL increase in WMHV (CI: 1.01, 1.43). Finally, we observed that every unit reduction on the Word Definition task was associated with an increase of 2.30 mL in ventricular volume (CI: 0.31, 4.30).When removing childhood cognitive abilities from this model, there was a reduction in effect size so that the association became non-significant (-1.17, CI:-3.01, 0.66). No significant associations were observed between the cognitive tasks and cross-sectional measures of whole brain and hippocampal volume. A full summary of the models’ results is reported in Supplementary table 4.

**Figure 5.**
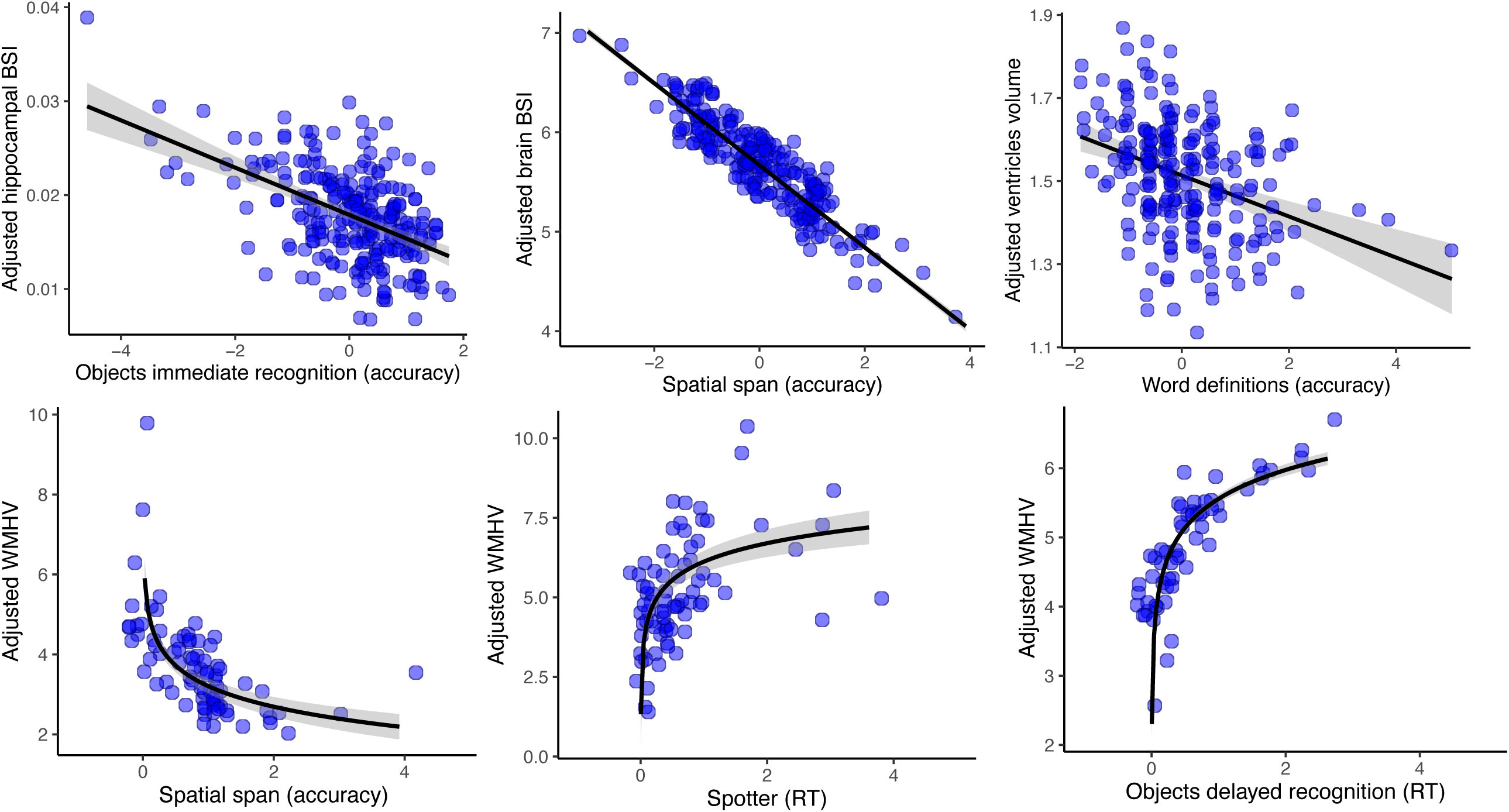
Scatter plots showing the associations between the Cognitron tasks and the biomarkers of AD, white matter pathology and neurodegeneration.

### 3.2 Validation against standard supervised neuropsychological assessments

The Cognitron composite score derived from the Cognitron task scores which predicted the biomarkers (Objects delayed and immediate recognition, Spatial span, Word definitions, and Spotter) explained 26.62% of the variance of the Cognitron tasks and showed a positive correlation with the total composite score of the standard assessments (rho=0.42, p<0.001), which explained 26.60% of the variance of the standard assessments, and the PACC (rho=0.32 p<0.001). Higher Cognitron memory composite score, accounting for 54.63% of the variance, significantly correlated with the standard assessments’ memory composite score, which explained 43.10% of the variance (rho=0.43, p<0.001) (Figure 6).

**Figure 6.**
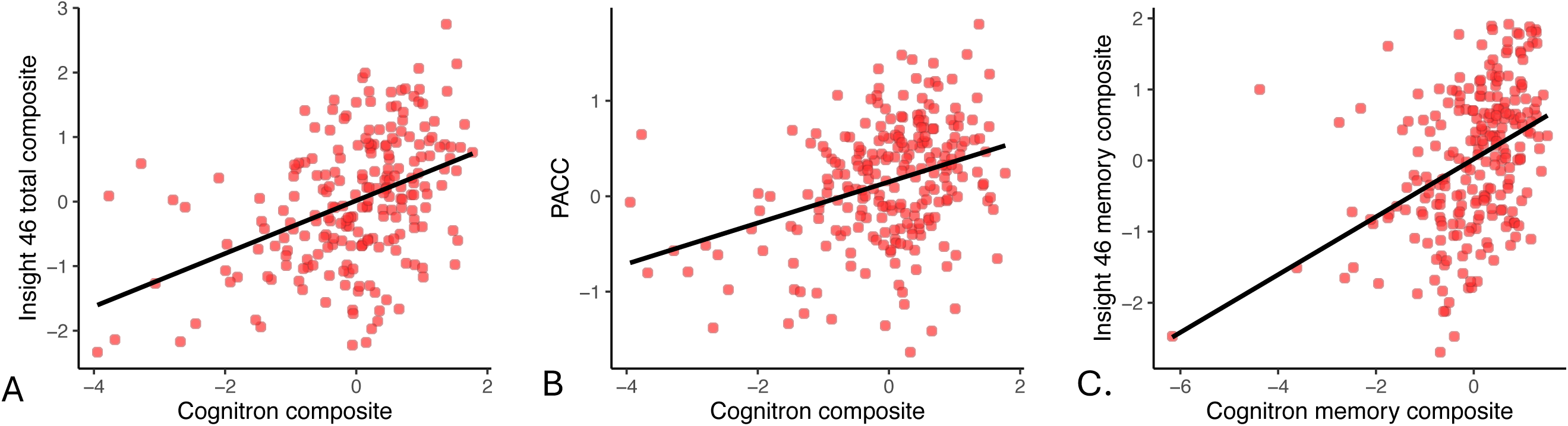
Plots showing the association between: (A) the Cognitron composite score generated from the tasks which predicted the imaging biomarkers and the Insight 46 total composite score (B) the Cognitron composite score generated from the tasks which predicted the imaging biomarkers and the PACC (C) the Cognitron memory composite score and the Insight 46

## 4 Discussion

In this study, we remotely deployed a battery of computerised tasks to the Online 46 cohort, a population-based cohort of individuals born in England, Scotland, and Wales within the same week of March 1946. We identified the tasks that predicted biomarkers of AD and neurodegeneration and verified that these correlated with standard supervised cognitive assessments.

The key finding of this study is that Aβ-positive participants were significantly slower than Aβ-negative participants on the Objects delayed recognition task. Lower accuracy on the same task was also associated with amyloid positivity, although approaching significance with a weaker effect. These findings were further supported when examining the relationship between cognitive performance and amyloid deposition measured as a continuous variable (i.e., SUVR). Notably, we found that impairments in the Aβ-positive group affected delayed rather than immediate memory recognition, likely reflecting a process of accelerated forgetting. Previous studies measured this after 30 minutes and 7 days, showing effects in individuals with preclinical AD only after 7 days [47,48]. However, we detected this deficit at a single timepoint, which is advantageous since testing over multiple days is more challenging and often leads to lower compliance.

Previous studies have found slower RT and RT inter-individual variability in individuals with MCI and AD, as well as slower RT in attentional tasks for AD patients [49,50]. Additionally, lower performance on simple and choice reaction time tasks has been found to predict memory deficits in AD patients [49]. In line with these findings, we demonstrated that cognitive impairment in Aβ-positive individuals is more evident when examining the RT required to perform a memory task rather than accuracy alone. Notably, slower RT was significantly associated with poorer accuracy only in the Aβ-positive group, which accords with memory access being more effortful and therefore requiring more processing time [48]. This may reflect the early stages of an amnestic syndrome, where overt memory deficits are obscured by compensatory mechanisms, such as taking more time to complete a task. Therefore, RT as a measure of processing speed and as potential evidence of compensation for memory problems appears to be a good indicator of early AD-related cognitive changes. Studies should further investigate the relationship between RT and accuracy on memory tasks in individuals with or at risk of developing dementia.

Higher rates of whole brain and hippocampal atrophy between were associated with poorer performance on the Spatial span and Objects recognition tasks. The annual atrophy rate in these regions accelerates in individuals with MCI compared to healthy controls and is even higher in progressive compared to stable MCI [51,52]. Atrophy rate has been shown to be a good indicator of disease progression, correlating with cognitive decline and predicting conversion from MCI to AD [53,54]. Our findings support that these tasks hold value in the remote assessment of disease risk and progression. Future studies should focus on the longitudinal administration of the tasks to map cognitive impairment trajectories and examine their association with the accumulation of AD pathology. Conversely, in line with previous findings, we did not find any significant association between cognitive performance and cross-sectional brain volumes, indicating that longitudinal measurements of atrophy change may be a more sensitive measure of preclinical AD [55].

We also found that performance on the Word Definitions task, the main Cognitron measure of crystallised cognitive abilities, was associated with increased ventricular volume. Ventricular enlargement occurs naturally with age, but it can also predict conversion from MCI to AD and correlates with the degree of cognitive impairments in individuals with AD [56,57]. Late-life cognitive ageing typically affects fluid intelligence while sparing crystallised cognitive abilities, which are usually considered a better indicator for detecting early cognitive signs of AD [58]. Therefore, the association between Word Definitions and ventricular volume may reflect cognitive changes that deviate from the normal ageing trajectory. It is important to note that removing childhood cognitive abilities from the model resulted in a reduction in the estimated effect size and caused the relationship to lose statistical significance; this may reflect early life cognitive reserve having a protective effect on later-life cognitive deterioration.

In addition to the findings discussed above, the completion rate of the Cognitron battery suggests that remote computerised cognitive testing in elderly individuals at risk of developing dementia is both feasible and well-tolerated. Importantly, the completion rates of the tasks were not biased by the incidence of Aβ-positivity or by ApoE status, supporting the scope to use these cognitive tasks to measure cognitive abilities in preclinical AD.

There are however several limitations to this study. First, there is a difference in time between the data collection of the biomarkers and the standard cognitive tests and of the Cognitron tasks. Second, recruitment and participation to the Insight 46 study have been shown to be biased towards people with better health, higher education level and socioeconomic status, which may limit the generalisability of our sample, although this is common risk across any study cohort or epidemiological studies [59–61]. Additionally, there was variability in the number of participants who completed each task. Despite this, each task remained adequately powered, with a minimum of 229 subjects. Missing data might not be at random, as participants could not skip tasks, potentially excluding those with greater impairments. However, the inability to proceed with the assessment can itself serve as an indicator of cognitive status. Moreover, it is challenging to control the conditions under which the tests were completed remotely, a common issue for all remote computerised assessments. We attempted to control for this variability by including the device used to complete the assessment in the regression models, providing participants with instructions on the testing environment to adopt, and selecting tasks whose paradigm and design has relatively lower sensitivity to device differences. Finally, the usability of the Cognitron tasks is restricted to individuals with access to internet or a computerised device and a basic level of computer literacy. However, this limitation is becoming progressively less significant across generations. Furthermore, the tests can also be administered in-person for such individuals, with reduced need of resources compared to on-paper assessments and an automated scoring system.

Research investigating the use of computerised cognitive testing in the context of dementia diagnosis is growing significantly [62]. Among the tools which have been studied in relation to AD biomarkers, the C3PAD was able to discriminate Aβ-positive from Aβ-negative individuals who underwent Aβ-PET scans [63], while a digital version of the FNAME test has shown significant correlations with CSF levels of plasma pTau181 and Aβ42/40 ratio [64]. The CANTAB demonstrated to be able to identify individuals with AD and MCI, to detect cognitive changes of individuals with MCI longitudinally and to predict hippocampal volume and CSF biomarkers of T-tau, pTau181 and tau/Aβ42 ratio in individuals with MCI [65–67]. Cogstate, instead, showed sensitivity to longitudinal cognitive decline in Aβ- positive individuals, and a significant association with tau-PET and hippocampal atrophy [68,69]. However, these studies were conducted in supervised settings and have yet to the explore the remote applicability of the tests. Other studies have shown promise in testing elderly cognitively unimpaired individuals in unsupervised conditions and examined their cognitive abilities in relation to neurodegenerative biomarkers [70,71]. However, they have not investigated the administration of the tasks across multiple types of electronic devices or selected the most appropriate tasks from a broader set measuring different cognitive domains. In our study, we evaluated a comprehensive battery of tasks administered remotely in unsupervised conditions and run on a broad range of electronic devices. We identified a subset that can specifically target cognitive impairments associated with amyloid status and biomarkers indicative of AD severity and progression, while correlating with standard face-to-face assessments. Being easy to deploy and score while requiring minimal amount of time to complete, the Cognitron tasks represent a cost-efficient tool for large-scale screening of individuals at risk of AD, and potentially for monitoring patients who require longitudinal follow-ups, such as those with atypical presentation or who received a diagnosis at early stages of the disease [72]. This is particularly advantageous in clinical settings and for large cohorts, where the required resources for repeat assessments of patients are limited.

## Data Availability

Data sharing is subject to application through the National Survey of Health and Development portal (https://nshd.mrc.ac.uk)

## 5 Consent statement

The study was approved by the National Research Ethics Service Committee London (REC reference 19/LO/1774) and all participants provided written informed consent.

