## Supplementary table 1 for "Remote cognitive tests predict neurodegenerative biomarkers in the Insight 46 cohort"

Supplementary table 1. Details of the Insight 46 standard supervised cognitive assessments

|  | Test description | Cognitive domain | Scores analysed | References |
| --- | --- | --- | --- | --- |
| Logical memory (WMS-R) | Participants are read two stories composed by a series of simple but specific details and are asked to recall as many details as possible both immediately after hearing the story and after a set delay. | Memory immediate and delayed recall | Total correct | [37] |
| DSST (WAIS-R) | Participants are given a sheet with a series of digits and an empty box underneath each digit. The top of the sheet contains a key that pairs each of several digits with a corresponding symbol. The participant's task is to fill in the empty boxes with the corresponding symbol as quickly as possible. | Attention and processing speed | Total correct | [36] |
| FNAME-12* | Participants are shown 12 face-name pairs and are asked to recall the association immediately after the stimuli presentation and after a set delay. | Associative memory | Total correct (accelerated forgetting) | 19,38 |
| MMSE | This is a widely used tool which evaluates various aspects of cognitive status, including orientation, memory, attention, calculation, language, and visual-spatial skills | Overall cognitive abilities | Total score | [35] |
| Matrix reasoning (WASI) | Participants are shown incomplete patterns and are required to identify the missing piece from a set of options. | Non-verbal reasoning/ visuospatial abilities | Total correct | [40] |
| CRT | Participants are shown a cue arrow or word indicating right or left and must press the button that corresponds to the stimulus. | Processing speed | Mean RT for correct answers, error rate, intraindividual variability (std/mean) | [27,44] |
| Graded naming test | Participants are shown a series of objects in order of increasing difficulty and are asked to name them. | Language | Total correct | [43] |
| AMIPB - Complex figure drawing (7-day recall)* | Participants are asked to replicate a detailed geometric figure and to draw it from memory after a set amount of time. | Memory immediate and delayed recall | Figure copy, immediate recall, 30-minute recall, 7-day recall, accelerated forgetting (7-day recall/30-min recall) | [19,41] |
| PACC | This is a cognitive assessment tool designed to detect subtle cognitive deficits in individuals at the preclinical stage of AD. It includes the MMSE, the Logical Memory Delayed Recall, the Digit Symbol Substitution Test and the 12-item Face-Name Associative Memory Exam | Overall cognitive abilities | Total composite score | [39] |

WMS = Wechsler Memory Scale Revised, DSST = Digit symbol substitution test, FNAME-12 = 12-item Face-Name test, MMSE = Minimental State Examiner, WASI = Wechsler Abbreviated Scale of Intelligence, CRT = choice reaction time, AMIPB=Adult memory and information processing battery, PACC=Preclinical Alzheimer’s cognitive composite. * These measures were part of an ‘accelerated-forgetting’ assessment performed seven days after their research visit.
