## Supplementary table 2 for "Remote cognitive tests predict neurodegenerative biomarkers in the Insight 46 cohort"

Supplementary table 2. Participants completion rate for each Cognitron task.

|  | A*β*-positive  N=66 | A*β*-negative  N=175 | Total  N=255* |
| --- | --- | --- | --- |
| Objects immediate recognition | 66 (100%) | 175 (100%) | 255 (100%) |
| Motor control | 64 (97.0%) | 175 (100%) | 253 (99.2%) |
| Blocks | 63 (95.5%) | 173 (98.9%) | 250 (98.0%) |
| Digit span^†^ | 62 (93.9%) | 171 (97.7%) | 245 (96.1%) |
| Spatial span | 61 (92.4%) | 171 (97.7%) | 244 (95.7%) |
| Stroop | 60 (90.9%) | 171 (97,7%) | 243 (95.3%) |
| 2D Manipulations^†^ | 60 (90.9%) | 169 (96.6%) | 241(94.5%) |
| Word definitions | 60 (90.9%) | 170 (97.1%) | 242 (94.9%) |
| Verbal reasoning | 60 (90.9%) | 170 (97.1%) | 242 (94.9%) |
| Forager | 60 (90.9%) | 170 (97.1%) | 242 (94.9%) |
| Spotter^†^ | 59 (89.4%) | 168 (96.0%) | 239 (93.7%) |
| Objects delayed recognition | 60 (90.9%) | 168 (96.0%) | 240 (94.1%) |

* amyloid status missing for 14 participants

^†^ after removal of participants showing indicators of lack of compliance
