## Supplementary table 3 for "Remote cognitive tests predict neurodegenerative biomarkers in the Insight 46 cohort"

Supplementary table 3. Demographic differences between individuals who attempted and did not attempt the Cognitron battery

|  | Did not attempt tests | Attempted tests | Group differences |
| --- | --- | --- | --- |
| N | 122 | 274* |  |
| Sex, N (%) Male | 66 (54.10%) | 140 (51.09%) | X^2^_(1)_ =0.20, p=0.66 |
| Education N (%) |  |  | X^2^_(3)_ =5.17, p=0.16 |
| < 16 years | 67 (54.92%) | 47 (17.15%) |  |
| High School | 35 (28.69%) | 97 (35.40%) |  |
| Higher-level Degree | 20 (16.39%) | 60 (21.90%) |  |
| Handedness  N (%)  right-handed | 112 (91.80%) | 253 (92.33%) | X^2^_(1)_=-31, p=0.97 |
| Childhood cognitive ability mean (SD) | 0.27 (0.76%) | 0.46 (0.72%) | t_(665.44)_=-6.69, p<0.001, [CI:-0.38, 0.21] |
| Adult socioeconomic position †  N (%) |  |  | X^2^_(5)_=7.67, p=0.18 |
| Unskilled | 2 (1.64%) | 0 (0%) |  |
| Partly skilled | 4 (3.28%) | 13 (4.74%) |  |
| Skilled manual | 13 (10.66%) | 21 (7.66%) |  |
| Skilled nonmanual | 29 (23.77%) | 54 (19.71%) |  |
| Intermediate | 63 (51.64) | 151 (55.11%) |  |
| Professional | 11 (9.02%) | 35 (12.77%) |  |
| Amyloid positive N (%) | 36 (29.51%) | 73 (26.64%) | X^2^_(1)_ =0.13, p=0.72 |
| ApoE status ‡  N (%) e4 carriers | 41(33.61%) | 75 (27.37%) | X^2^_(2)_ =2.00, p=0.37 |
| MMSE  mean (SD) | 29.06 (0.98) | 29.42 (0.77) | W=13229, p<0.001 |
| PACC mean (SD) | -0.17(0.72) | 0.12 (0.62) | t_(205.76)_=-3.85, p<0.001 [CI: -0.44, -0.14] |

* missing amyloid status for 14 participants

† Derived from participant’s education at age 53

‡ From age 60 to age 64 and from age 69-71 if not known earlier
