## Supplementary table 1 for "Remote cognitive tests predict neurodegenerative biomarkers in the Insight 46 cohort"

Supplementary table 4. Results of the models conducted to look at the relationship between the Cognitron tasks and the biomarkers.

|  | SUVR^*^ | WMHV  (Log scale) ^†^ | Whole brain volume^*^ | Whole brain BSI^*^ | Hippocampal volume^*^ | Hippocampal BSI^*^ | Ventricular volume^*^ | Ventricular BSI^*^ |
| --- | --- | --- | --- | --- | --- | --- | --- | --- |
| Objects immediate recognition | -0.014  (CI: -0.049, 0.021) | -0.021  (CI: -0.165,0.113) | 4.435  (CI: -1.336, 10.205) | -0.35  (CI: -0.746, 0.046) | 0.076  (CI: -0.001, 0.153) | -0.006  (CI: -0.012, -0.001) | 0.52  (CI: -1.325, 2.366) | -0.079  (CI: -0.19, 0.033) |
| Motor control | 0.008  (CI: -0.028, 0.045) | 0.087  (CI: -0.059, 0.25) | -7.433  (CI: -13.534, -1.333) | -0.04  (CI: -0.468, 0.389) | -0.069  (CI: -0.153, 0.015) | -0.003  (CI: -0.009, 0.003) | -0.403  (CI: -2.409, 1.604) | -0.073  (CI: -0.194, 0.047) |
| Blocks | -0.022  (CI: -0.054, 0.01) | 0.02  (CI: -0.119, 0.156) | 0.238  (CI: -5.209, 5.685) | -0.254  (CI: -0.627, 0.118) | -0.029  (CI: -0.103, 0.045) | 0.000  (CI: -0.005, 0.005) | -0.701  (CI: -2.44, 1.039) | -0.058  (CI: -0.16, 0.044) |
| Digit span | 0.026  (CI: -0.018, 0.07) | -0.039  (CI: -0.213, 0.162) | 4.622  (CI: -2.889, 12.133) | -0.105  (CI: -0.608, 0.398) | -0.011  (CI: -0.113, 0.091) | 0.000  (CI: -0.006, 0.007) | 0.531  (CI: -1.873, 2.936) | -0.046  (CI: -0.184, 0.093) |
| Spatial Span | -0.009  (CI: -0.042, 0.024) | -0.191  (CI: -0.333, -0.046) | 4.177  (CI: -1.35, 9.704) | -0.42  (CI: -0.795, -0.046) | 0.043  (CI: -0.034, 0.119) | -0.003  (CI: -0.008, 0.002) | -1.254  (CI: -3.058, 0.55) | -0.086  (CI: -0.189, 0.017) |
| Stroop | 0.002  (CI: -0.035, 0.038) | -0.006  (CI: -0.165, 0.155) | 3.058  (CI: -3.112, 9.228) | -0.194  (CI: -0.617, 0.229) | -0.028  (CI: -0.113, 0.057) | -0.003  (CI: -0.009, 0.003) | -0.81  (CI: -2.826, 1.206) | -0.064  (CI: -0.18, 0.051) |
| 2D Manipulations | 0.007  (CI: -0.029, 0.043) | -0.098  (CI: -0.262, 0.067) | 2.727  (CI: -3.29, 8.745) | 0.169  (CI: -0.243, 0.581) | 0.041  (CI: -0.042, 0.124) | 0.001  (CI: -0.004, 0.007) | -0.828  (CI: -2.773, 1.118) | 0.005  (CI: -0.108, 0.117) |
| Word definitions | -0.011  (CI: -0.048, 0.027) | -0.1  (CI: -0.259, 0.065) | 2.334  (CI: -3.902, 8.571) | 0.024  (CI: -0.401, 0.449) | 0.057  (CI: -0.029, 0.143) | -0.004  (CI: -0.009, 0.002) | -2.301  (CI: -4.295, -0.307) | -0.042  (CI: -0.158, 0.074) |
| Verbal reasoning | -0.002  (CI: -0.038, 0.034) | 0.026  (CI: -0.124, 0.169) | -0.824  (CI: -6.856, 5.209) | -0.209  (CI: -0.617, 0.198) | -0.037  (CI: -0.12, 0.046) | 0.000  (CI: -0.006, 0.005) | 1.107  (CI: -0.839, 3.052) | -0.007  (CI: -0.119, 0.104) |
| Forager | -0.009  (CI: -0.043, 0.025) | -0.035  (CI: -0.198, 0.126) | 4.008  (CI: -1.727, 9.742) | -0.184  (CI: -0.577, 0.209) | 0.031  (CI: -0.049, 0.11) | -0.002  (CI: -0.007, 0.003) | 0.185  (CI: -1.679, 2.048) | 0  (CI: -0.108, 0.107) |
| Spotter | 0.018  (CI: -0.027, 0.063) | -0.01  (CI: -0.295, 0.216) | 5.582  (CI: -2.108, 13.272) | -0.464  (CI: -0.98, 0.052) | 0.033  (CI: -0.074, 0.14) | 0.006  (CI: -0.001, 0.012) | -0.481  (CI: -2.978, 2.015) | -0.028  (CI: -0.171, 0.115) |
| Objects delayed recognition | -0.037  (CI: -0.072, -0.003) | 0.027  (CI: -0.132, 0.181) | 3.641  (CI: -2.193, 9.475) | -0.319  (CI: -0.718, 0.079) | 0.054  (CI: -0.027, 0.134) | -0.004  (CI: -0.01, 0.001) | 0.071  (CI: -1.825, 1.966) | -0.061  (CI: -0.17, 0.049) |
| Objects immediate recognition RT | 0.004  (CI: -0.021, 0.029) | 0.093  (CI: -0.03, 0.261) | -2.965  (CI: -7.127, 1.198) | 0.174  (CI: -0.112, 0.459) | 0.013  (CI: -0.043, 0.069) | 0.001  (CI: -0.003, 0.005) | 0.971  (CI: -0.353, 2.295) | 0.043  (CI: -0.037, 0.123) |
| Motor control RT | -0.001  (CI: -0.025, 0.022) | 0.04  (CI: -0.067, 0.211) | -0.665  (CI: -4.687, 3.357) | -0.158  (CI: -0.436, 0.119) | -0.021  (CI: -0.076, 0.034) | -0.002  (CI: -0.006, 0.001) | -0.321  (CI: -1.626, 0.983) | -0.027  (CI: -0.105, 0.052) |
| Blocks RT | -0.016  (CI: -0.052, 0.021) | 0.127  (CI: -0.029, 0.291) | -0.966  (CI: -7.282, 5.349) | -0.062  (CI: -0.484, 0.359) | -0.052  (CI: -0.137, 0.034) | -0.002  (CI: -0.008, 0.004) | 0.211  (CI: -1.809, 2.232) | 0.003  (CI: -0.113, 0.118) |
| Digit span RT | 0.021  (CI: -0.023, 0.065) | 0.078  (CI: -0.101, 0.277) | 3.426  (CI: -4.135, 10.986) | -0.388  (CI: -0.888, 0.113) | 0.027  (CI: -0.075, 0.129) | -0.001  (CI: -0.008, 0.006) | 0.157  (CI: -2.26, 2.575) | -0.074  (CI: -0.213, 0.064) |
| Spatial Span RT | 0.008  (CI: -0.031, 0.046) | 0.148  (CI: -0.001, 0.312) | -3.077  (CI: -9.563, 3.408) | -0.173  (CI: -0.619, 0.272) | -0.036  (CI: -0.125, 0.054) | 0.000  (CI: -0.006, 0.005) | -0.712  (CI: -2.829, 1.406) | -0.064  (CI: -0.186, 0.057) |
| Stroop RT | -0.003  (CI: -0.034, 0.027) | 0.009  (CI: -0.119, 0.152) | -2.538  (CI: -7.601, 2.525) | 0.197  (CI: -0.148, 0.541) | 0.028  (CI: -0.041, 0.098) | -0.001  (CI: -0.005, 0.004) | 0.25  (CI: -1.406, 1.907) | 0.044  (CI: -0.05, 0.138) |
| 2D Manipulations RT | -0.022  (CI: -0.056, 0.013) | 0.042  (CI: -0.113, 0.207) | 1.113  (CI: -4.714, 6.941) | -0.191  (CI: -0.599, 0.217) | -0.043  (CI: -0.123, 0.038) | -0.002  (CI: -0.007, 0.004) | 0.673  (CI: -1.209, 2.555) | -0.011  (CI: -0.123, 0.101) |
| Word definitions RT | 0.007  (CI: -0.027, 0.041) | 0.064  (CI: -0.095, 0.229) | 2.067  (CI: -3.677, 7.811) | 0.032  (CI: -0.359, 0.422) | 0.06  (CI: -0.019, 0.139) | 0.002  (CI: -0.004, 0.007) | 0.306  (CI: -1.554, 2.166) | 0.032  (CI: -0.075, 0.139) |
| Verbal reasoning RT | -0.01  (CI: -0.043, 0.023) | -0.042  (CI: -0.174, 0.097) | -2.4  (CI: -7.866, 3.067) | 0.004  (CI: -0.371, 0.38) | -0.024  (CI: -0.099, 0.052) | -0.001  (CI: -0.006, 0.004) | 0.55  (CI: -1.22, 2.32) | -0.036  (CI: -0.138, 0.067) |
| Forager RT | 0.014  (CI: -0.04, 0.06) | 0.1  (CI: -0.14, 0.34) | -6.87  (CI: -15.59, 1.84) | 0.127  (CI: -0.47, 0.72) | -0.09  (CI: -0.21, 0.03) | 0.002  (CI: -0.01, 0.01) | 0.9  (CI: -1.94, 3.73) | -0.012  (CI: -0.176, 0.15) |
| Spotter RT | 0.004  (CI: -0.033, 0.041) | 0.177  (CI: 0.005, 0.36) | -5.306  (CI: -11.619, 1.007) | 0.383  (CI: -0.043, 0.809) | -0.044  (CI: -0.131, 0.044) | 0.001  (CI: -0.004, 0.007) | 1.814  (CI: -0.224, 3.851) | 0.053  (CI: -0.065, 0.171) |
| Objects delayed recognition RT | 0.053  (CI: 0.01, 0.095) | 0.237  (CI: 0.035, 0.453) | -1.841  (CI: -9.144, 5.462) | 0.105  (CI: -0.391, 0.6) | -0.025  (CI: -0.126, 0.075) | -0.002  (CI: -0.009, 0.004) | 0.302  (CI: -2.063, 2.667) | -0.014  (CI: -0.149, 0.121) |

RT=response time, WMHV = white matter hyperintensity volume, CI=confidence interval, SUVR = standard uptake volume ratio, BSI = boundary shift integral, *= linear regression , †= generalised linear model with gamma log link function,
